# The SLaM Brain Health Clinic: a remote biomarker enhanced memory clinic for patients with mild cognitive impairment within an NHS mental health trust

**DOI:** 10.1101/2024.04.23.24303268

**Authors:** Ashwin V. Venkataraman, Pooja Kandangwa, Roos Lemmen, Rutvi Savla, Mazda Beigi, Devon Hammond, Daniel Harwood, Justin Sauer, Latha Velayudhan, Clive Ballard, Anna-Katharine Brem, Chris Kalafatis, Dag Aarsland

## Abstract

**Background:** The novel South London and Maudsley Brain Health Clinic (SLaM BHC) leverages advances in remote consultations and biomarkers to provide a timely, cost-efficient and accurate diagnosis in mild cognitive impairment (MCI).

**Aims:** To describe the organisation, patient cohort, and acceptability of the remote diagnostic and interventional procedures.

**Method:** We describe the recruitment, consultation setup, the clinical and biomarker program, and the two online group interventions for cognitive wellbeing and lifestyle change. We evaluate the acceptability of the remote consultations, lumbar puncture (LP), saliva genotyping and remote cognitive and functional assessments.

**Results:** We present the results of the first 68 (mean age 73, 55% female, 43% ethnic minority) of 146 patients who enrolled for full remote clinical, cognitive, genetic, cerebrospinal fluid, and neuroimaging phenotyping. 86% were very satisfied/ satisfied with the remote service. 67% consented to LP and 95% of those were very satisfied, all having no significant complications. 93% found taking saliva genotyping very easy/easy and 93% found the cognitive assessments instructions clear. 98% were satisfied with the cognitive wellbeing groups and 90% of goals were achieved in the lifestyle intervention group.

**Conclusions:** The SLaM BHC provides a highly acceptable and safe clinical model for remote assessments and lumbar punctures in a representative, ethnically diverse population. This allows early and accurate diagnosis of Alzheimer’s, differentiation from other MCI causes and targets modifiable risk factors. This is crucial for future disease modification, ensuring equitable access to research, and provides precise, timely and cost-efficient diagnoses in UK mental health services.

## INTRODUCTION

Dementia, of which the largest cause is Alzheimer’s Disease (AD), affects 50 million people globally with a predicted three-fold increase by 2050. In the UK, revised increased estimates suggest 1.7 million people will have dementia by 2040 (1). An even higher number of people have MCI, many of whom are in the prodromal stage of AD (2). The emergence of new disease-modifying therapies for AD is a huge opportunity, but also a challenge (3–5). 30,200 people per year are expected to be eligible for disease modifying monoclonal antibody therapies for AD in the UK (6) with expected wait times for access forecast to be 56 months in 2023, increasing to 129 months in 2029, hence the need for rapid change and innovative approaches in this space (7,8).

There are now rapid advances in digital, imaging, and molecular biomarkers of AD (9– 13), remote assessment opportunities (14,15), alongside the emergence of new therapies and knowledge of targeting modifiable risk factors (16). Early accurate aetiological diagnosis of AD is crucial to enable adequate treatment and is in line with public attitudes (17), but the uptake of the diagnostic biomarkers is extremely low in some countries, including the UK (18). There is therefore a clear need for memory services to rapidly adapt to this new landscape for greater patient benefit, and to match the molecular and digital biomarker developments globally in this field, (19). Notably there is a huge gap that exists between the demand and assessment - 99% of people with MCI never receive a diagnosis and are not referred to memory clinics, and this capacity must increase (20). This is particularly important in mental health trusts in the UK who see 92% of patients with memory complaints with the remainder seen by geriatrics and neurology (18).

The SLaM BHC is an innovative, remote service within a mental health trust that leverages advances in accessibility of remote consultations combined with detailed biomarker assessment with the aim to address these new challenges for the healthcare system. Here we describe the organisation, the diagnostic and intervention procedures, and the interventional groups. We describe the key characteristics of the first 146 referrals to the SLaM BHC, and the experience, feasibility and acceptability, of those signed up for the linked BHC research project.

## METHODS

### 1 Recruitment and participants

The South London and Maudsley NHS Foundation Trust in the UK covers a catchment population of over 1.3 million people across four London boroughs. Within this trust referrals to the Brain Health Clinic were made via three memory services (Croydon, Lewisham, and the combined service for Southwark and Lambeth) after an initial clinical assessment with possible additional brain imaging.

The SLaM BHC research protocol was approved under the Research Ethics Committee (REC) 22/SC/0109 (South Central - Berkshire B, UK), was registered on ClinicalTrials.gov ID NCT06379594, and enabled the use of CSF, genetic and remote cognitive and functional biomarkers for all research participants. Out of the total 5751 referrals for all cognitive problems to the three memory services, 146 referrals were accepted to SLaM BHC and fulfilled inclusion/exclusion criteria below. The clinic began taking referrals from three SLaM memory services in October 2021 and closed to referrals in July 2023, and continued seeing the research participants. Up to 1st January 2024, 68 have consented and completed the initial assessment of the SLaM BHC research project, 40 patients will be approached during 2024, and 45 declined or were ineligible and were of similar demographics to those that consented.

Inclusion criteria for the SLaM BHC, and the research project, were patients referred by SLaM memory clinics either with a formal diagnosis of MCI, subjective cognitive impairment, or mild dementia when the case was aetiologically complex. Additional inclusion criteria were the ability to access the clinic via telephone or video conferencing. Exclusion criteria were a diagnosis of moderate-severe dementia, or those unwilling or unable to provide written consent. All medications and treatments were permitted concurrently whilst engaging in this study and were flagged at the time of referral if affecting cognition. Figure 1 below shows an overview of the SLaM BHC.

**Figure 1:**
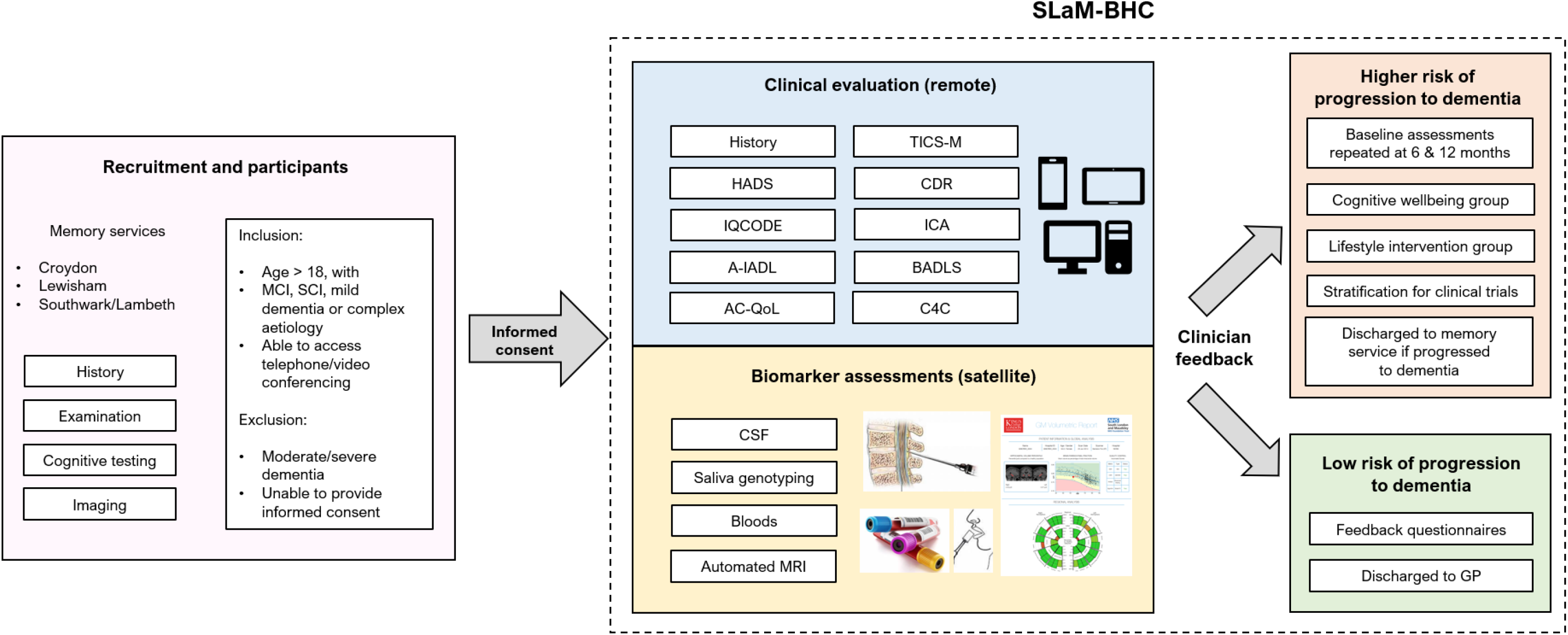
Overview of the SLaM BHC showing the recruitment and participants from 3 memory services, inclusion criteria including those with mild cognitive impairment (MCI) and subjective cognitive impairment (SCI), or mild dementia of uncertain or complex aetiology and exclusion criteria following referral to the SLaM BHC. Clinical evaluation comprised of history, the Hospital Anxiety and Depression Scale (HADS), Informant Questionnaire on Cognitive Decline in the Elderly (IQ-CODE), Amsterdam iADL functional assessment (A-IADL), Adult Carer Quality of Life Questionnaire (AC-QoL), Telephone Interview for Cognitive Status for Memory (TICS-M), Integrated Cognitive Assessment (ICA) and patient reported experience and outcome measures, Bristol Activities of Daily Living Scale (BADLS), and assessment for Consent for Contact for research (C4C). Satellite procedures for biomarker assessments included lumbar puncture for CSF, saliva genotyping, bloods, and automated MRI. Following this information individuals were stratified into higher risk of progression to dementia or lower risk of progression to dementia with listed outcomes below, with all followed up after 6 and 12 months under the research component.

### 2 The consultation setup

All patients seen in the Brain Health Clinic underwent remote assessments using virtual conferencing (via MS Teams), with appropriate help from a caregiver/family member when needed, by a psychiatrist or an experienced nurse clinician with opportunities for support from the SLaM Digital inclusion team (21). In some cases only telephone assessment was possible. Remote clinical evaluation and satellite biomarker assessments were performed with individual feedback to patients and families via telephone/virtual conferencing following consensus diagnosis of the stage and aetiology in a virtual MDT.

### 3 The clinical evaluation program

The current remote baseline assessment protocol included a detailed patient history, the Telephone Interview for Cognitive Status (TICS), Clinical Disease Rating (CDR), the Hospital Anxiety and Depression Scale (HADS), Consent for Contact for research (C4C) and patient reported experience and outcome measures. The Informant Questionnaire on Cognitive, Decline in the Elderly (IQ-CODE), digital version of Amsterdam iADL functional assessment, Bristol Activities of Daily Living Scale (BADLS) and computerised cognitive assessments were emailed and sent to patients. The Integrated Cognitive Assessment (ICA) is a 5-minute computerised cognitive test based on a rapid categorization task that employs an artificial intelligence model to improve its accuracy in detecting cognitive impairment (22). The ICA is self-administered and independent of language (23,24). We will focus on the feasibility aspects and not present the individual results of these tests.

### 4 The biomarker program

The BHC research project biomarker programme for those consenting included CSF for AD markers, saliva for genetics. In addition bloods for AD markers, and MRI are included but not presented here. Lumbar puncture was performed by a neurologist on a pay-for-service basis at the BRC Clinical Research Facility. CSF analysis was performed using the ElectroChemiLuminescence Immunoassay Instrument: Cobas® 6000 analyzer series. The Assays are: Elecsys® β-Amyloid(1-42) CSF, Elecsys® Phospho-Tau (181P) CSF & Elecsys® Total-Tau CSF) at a private local laboratory. Saliva samples were analysed by Cytox Group Limited, employing a polygenic risk scoring algorithm, genoSCORE™LAB, including APOE genotyping, to identify those at highest genetic risk of AD using genetic data from the saliva sample (25). Additionally, participants provided a blood plasma sample for use in future diagnostic dementia biomarker studies (13,26). Automated volumetric MRI analysis pipelines extracted regional volumes compared to normative populations using a geodesic information flow algorithm in addition to training on SLaM Image Bank (11,27,28) following routine structural MRI acquisition as per dementia scanning protocols.

Patients who were diagnosed with dementia while under follow up at the SLaM BHC were transferred back to the memory service. Patients who were not diagnosed with a neurodegenerative disease following assessment by the SLaM BHC were discharged to their GP. All were followed up under the research component at the 6 and 12 month time point regardless of risk.

### 5. The two intervention groups

The SLaM BHC developed two fully remote, teams-based, psychological intervention groups as part of the clinical procedure that all patients were invited for. The first was the Cognitive Wellbeing Group which focussed on psychoeducation on brain anatomy, cognition, MCI, dementia, and psychological concepts with strategies to manage memory and mood-related difficulties. The second group was the Lifestyle Intervention Group which focussed on dementia prevention and the impact of lifestyle factors on cognition and the potential for lifestyle changes, including goal setting, physical health, physical activity, nutrition, sleep, keeping your mind active, social activity and compensatory techniques for memory. Both groups consisted of 6 to 8 patients, ran over eight one-hour sessions per round and was led by two clinicians, one psychologist and one psychology assistant.

### 6 Feasibility and acceptability assessments

Participant feedback was analysed with a semi-structured interview outcome. Feedback on the clinic and individual virtual technologies were given specifically for lumbar punctures, genoscore, and the patient reported experience and outcome measures, digital biomarkers (Amsterdam iADL, ICA), feedback questionnaires, alongside semi-structured interviews for the groups.

## RESULTS

### The cohort and feasibility

As seen in Table 1, the full cohort is fairly representative for an NHS memory clinic (18) with regards to age (mean 75y) and gender (64% female) with a higher percentage of ethnic diversity (58% white, 42% ethnic minority) and lower education (53% having secondary school or less). Of 135 patients the majority, 73%, were able to complete the virtual assessment, whereas 27% could do telephone assessment only. The majority of participants were able to complete the full clinical, cognitive assessments and the biomarker acquisition procedures.

**Table 1:** Demographic and clinical characteristics of the cohort stratified by number of patients completing the various biomarker procedures.

|  | Total cohort | Consented to research study | Consented to lumbar puncture | Cognitive Wellbeing Group | Lifestyle management group |
| --- | --- | --- | --- | --- | --- |
| **n** | 146 | 68 | 45 | 25 | 4 |
| **Age** (mean, range) | 75 (53 - 96) | 73 (53 – 89) | 71 (53 - 89) | 73 (53 – 89) | 59 (53 – 64) |
| **Gender** (n,%) |  |  |  |  |  |
| Female | 94 (64) | 37 (55) | 26 (58) | 17 (68) | 4 (100) |
| Male | 52 (36) | 31 (45) | 19 (42) | 8 (32) | 0 |
| **Ethnicity** (n,%) |  |  |  |  |  |
| Asian | 15 (10) | 10 (15) | 7 (15) | 3 (12) | 0 |
| Black | 33 (23) | 14 (21) | 8 (18) | 7 (28) | 2 (50) |
| Mixed ethnicity | 9 (6) | 1 (1) | 0 | 1 (4) | 0 |
| White | 85 (58) | 39 (57) | 27 (60) | 14 (56) | 2 (50) |
| Other ethnic group | 4 (3) | 4 (6) | 3 (7) | 0 | 0 |
| **Highest level of education** (n,%) |  |  |  |  |  |
| Primary school | 12 (8) | 2 (3) | 1 (2) | 0 | 0 |
| Secondary school | 65 (45) | 35 (51) | 27 (60) | 13 (52) | 3 (75) |
| Bachelor's degree or equivalent | 30 (21) | 17 (25) | 9 (20) | 8 (32) | 0 |
| Master's degree or equivalent | 3 (2) | 6 (9) | 5 (11) | 1 (4) | 1 (25) |
| PhD, doctorate or equivalent | 12 (8) | 7 (10) | 3 (7) | 2 (8) | 0 |
| Missing | 24 (16) | 1 (2) | 0 | 1 (4) | 0 |
| **Cognitive stage** (n,%) |  |  |  |  |  |
| MCI | 114 (78) | 56 (82) |  |  |  |
| Dementia | 27 (19) | 10 (15) |  |  |  |
| Other | 5 (3) | 2 (3) |  |  |  |

Of the 68 who consented to the research protocol, 45 (66%) also consented to LP. They had similar demographics to the full cohort. Of the 45 available, 26 patients have had an LP to date with 3 failed LPs. 15/23 available results (65%) had an Aβ42 value below the cut-off with 55% having a positive tTau/Aβ42 ratio. The average turn around time from CSF sample taken to result was 1 day (range 0-4 days).The median time from consent to CSF results back was 60 days. Of available Genoscore results for 35 patients, 18 (52%) had at least one e4 allele and 17 (48% has no e4 allele), with 19 (54%) having a high risk of progression to AD from the polygenic risk score, 6 (17%) being medium risk, and 10 (29%) being low risk. The average turnaround of genoscore results was 72 days (range 9-229 days). 25 participants attended the cognitive wellbeing group and 4 attended the lifestyle management group.

30% of patients were not diagnosed with a neurodegenerative disease following assessment and discharged to their GP. Cognitive staging of the full cohort showed 114 (78%) had MCI, and 27 (19%) fulfilled criteria for dementia, with similar proportions in the research cohort with 56 (82%) having MCI, and 10 (15%) having dementia at the last recorded time point (Table 1).

### Feedback

43 patients’ feedback for the overall remote BHC procedures were available and representative of the total and research cohort (mean age 74, 53% female, 52% ethnic minority). 17 (40%) found technologies for assessments and appointments either very easy or easy, 20 (47%) were neutral, and 3 (7%) found it difficult/very difficult and 3 (7%) did not respond. 26 (60%) of patients would recommend this to friends and family, 3 (7%) would not recommend it, and 11 (26%) did not respond. 30 (70%) were able to contact a team clinician when needed, 2 (5%) were not, and 7 (16%) did not respond. 37 (86%) patients were either very satisfied or satisfied with the overall service, 4 (9%) were neutral, 0 were dissatisfied, and 2 (5%) did not respond. Further details of feedback are provided Appendix Table 1.

For the LP procedure, 20 of 21 (95%) respondents were “very satisfied”, one (5%) satisfied”. 5 had had concerns prior to the procedure, all responded that they had had the opportunity to ask questions and thought the information sheet was helpful, and were able to contact a clinician when they needed, and only one (5%) had experienced complications (“anxiety about the results”) whereas 20 (95%) reported no complications (Appendix Table 2). Of the 45 genoscore feedback results, 42 (93%) found taking the saliva sample very easy or easy, 100% found the instructions clear, with 41 (91%) stating after taking it they would not have preferred doing this in clinic (Appendix Table 3).

45 participants completed the ICA feedback with 42 (93%) finding the instructions clear and 31 (69%) did not require support when completing the test (Appendix Table 4). Feedback on the intervention groups were available for 15 participants. As seen in Appendix Table 5, the feedback was very positive, with 14 (98%) finding the group very helpful and 1 (5%) neutral, and none unhelpful. All participants felt the groups helped them better understand both MCI and the impact of mental health on cognition. They were representative of the demographics of the whole cohort (Table 1). In the lifestyle intervention group 90% of goals that were set were achieved successfully.

## DISCUSSION

The SLaM BHC successfully provides an early and accurate diagnoses of AD in people with MCI, along with a safe and acceptable care model for various remote clinical, cognitive and biomarker assessments within an NHS mental health memory setting. This is crucial both in preparing for disease modification, stratifying risk and enhancing clinical research access with the opportunity for secondary prevention of cognitive decline.

The clinic has a number of strengths and demonstrated that it is now possible to provide remote clinical assessments for patients with high acceptability and very positive patient feedback. We also show that satellite in clinic biomarker evaluations for CSF, genotyping, bloods and neuroimaging are not only possible but highly acceptable with relatively fast turnaround times to the results once taken. Furthermore, we show this is possible in an ethnically diverse and representative cohort in South London with a higher proportion from less educated and more deprived backgrounds. We were able to show early and accurate diagnoses of AD in half of patients, with a third being discharged to the GP with no evidence of neurodegenerative disease. Finally we were able to implement effective secondary prevention interventions from the Cognitive Wellbeing Group and Lifestyle Management Group for elderly patients in the comfort of their own homes.

We know that older people are at higher risk of reduced physical and social activity, loneliness and depression which are all factors associated with more rapid cognitive and functional decline (29). Recent technological advances of remote memory assessments can provide an opportunity to re-evaluate how existing methods can be adapted for remote assessment and how digital technology can be used to automate cognitive assessments and data collection. In addition remote biomarkers provide the opportunity to further increase capacity and meet unmet demand. This is particularly important given most people with MCI never receive a diagnosis, and therefore there would need to be a necessary shift for accurate primary care based AD diagnoses using new methods to facilitate this (30).

The main limitation of the SLaM BHC to date is the small sample size. However, this is mainly due to limited capacity to include all eligible and consenting participants. In addition those referred to the service were potentially those more likely to engage in the program. Importantly, the participants in the research component did not differ from the overall referral cohort regarding age, ethnicity and education showing that it is representative. While we have shown the majority of this cohort were able to perform the procedures, a considerable proportion did not complete the digital set up. Digital exclusion is therefore a critical issue. This is expected to gradually become a smaller problem as digital competence increases. Mitigating strategies to support people who need this such as what was available from the SLaM Digital Inclusion Team (21) would help with this however this was only available for a limited time period.

Future plans for the SLaM BHC are related to scaling up assessments across other memory clinics in South London more widely and clinical workflows that are focused on the importance of actionable guidance towards prevention (31). The potential of using remote assessments and risk reduction that can be done in people’s homes and funnelled to GPs such as in the PREDICTOM study (30) and AD-RIDDLE (32), have the potential to improve the precision of referrals (33). Future blood based markers (13) and novel cognitive training games with additional clinical decision support tools may also be utilised in this remote diagnostic and interventional pathway.

## CONCLUSION

We have successfully shown that the remote SLaM BHC can provide an early and accurate diagnosis of AD in people with MCI in an NHS mental health trust in a diverse and representative population. It also provides an opportunity for addressing modifiable risk factors, provides safe and acceptable care for patients undergoing lumbar puncture and genotyping, and provides an acceptable model for remote assessments to increase the diagnostic capacity to meet unmet demand. This will be crucial in preparing for the prospect of disease modification, enhancing access disparities to clinical research and trials, alongside providing more precise diagnoses to patients and families.

## Declaration of interests

The authors declare there is no conflict of interest.

AVV has received grants from the Alzheimer’s Society, Alzheimer’s Research UK, and NIHR BRC, including an NIHR BRC Maudsley Neuroimaging Grant. DA has received research support and/or honoraria from Astra-Zeneca, H. Lundbeck, Novartis Pharmaceuticals, Biogen, and GE Health, and served as paid consultant for H. Lundbeck, Eisai, Heptares, Mentis Cura, Roche Diagnostics and Eli Lilly.

## Data Availability

All data produced in the present study are available upon reasonable request to the authors

## Acknowledgements

AVV is funded by the National Institute for Health Research (NIHR) as NIHR Clinical Lecturer and supported by the NIHR Maudsley Biomedical Research Centre at South London and Maudsley NHS Foundation Trust and King’s College London. The views expressed are those of the authors and not necessarily those of the NIHR or the Department of Health and Social Care. The SLaM BHC has received funding from Roche and the Maudsley Charity.

## APPENDIX

**Appendix Table 1:** Feedback on remote assessments in the SLaM BHC.

|  |  |
| --- | --- |
| Total | n = 43<br>n (%) |
| **1) How did you find using the technologies for your assessments and appointments?**<br>a. Easy/Very Easy<br>b. Neither<br>c. Difficult/Very difficult<br>d. Did not respond | 17 (40)<br>20 (47)<br>3 (7)<br>3 (7) |
| **2) Having completed the online assessments, would you recommend them to your friends and family?**<br>a. Yes<br>b. No<br>c. Not applicable<br>d. Did not respond | 26 (60)<br>3 (7)<br>3 (7)<br>11 (26) |
| **3) Were you able to contact a team clinician when you needed to?**<br>a. Yes<br>b. No<br>c. Not applicable<br>d. Did not respond | 30 (70)<br>2 (5)<br>4 (9)<br>7 (16) |
| **4) How satisfied are you overall with the service you have received?**<br>a. Satisfied/Very Satisfied<br>b. Neither satisfied or dissatisfied<br>c. Dissatisfied/Very Dissatisfied<br>d. Did not respond | 37 (86)<br>4 (9)<br>0<br>2 (5) |

**Appendix Table 2:** Lumbar puncture feedback.

|  |  |
| --- | --- |
| Total | n = 21<br>n (%) |
| **1) How satisfied were you with the procedure?**<br>a. Very satisfied<br>b. Satisfied<br>c. Neither satisfied/dissatisfied<br>d. Dissatisfied | 20 (95)<br>1 (5)<br>0 (0)<br>0 (0) |
| **2) Did you have any concerns about the procedure?**<br>a. Yes<br>b. No | 5 (24)<br>16 (76) |
| **3) Did you have an opportunity to ask questions**<br>a. Yes<br>b. No | 21(100)<br>0(0) |
| **4) Was the information sheet helpful?**<br>a. Yes<br>b. No | 21(100)<br>0(0) |
| **5) Did you experience any complications after the procedure?**<br>a. Yes<br>b. No | 1(5) "felt anxious about results"<br>20(95) |
| **6) Were you able to contact a clinician if needed**<br>a. Yes<br>b. No | 21(100)<br>0(0) |

**Appendix Table 3:** Genoscore feedback.

|  |  |
| --- | --- |
| Total | 45<br>n (%) |
| **1) How did you find taking the sample?**<br>a. Very Easy<br>b. Easy<br>c. Neither difficult nor easy<br>d. Difficult<br>e. Very Difficult | 17 (37)<br>25 (55)<br>3 (7)<br>0 (0)<br>0 (0) |
| **2) How did you find sending the sample?**<br>a. Very Easy<br>b. Easy<br>c. Neither difficult nor easy<br>d. Difficult<br>e. Very Difficult | 20 (44)<br>20 (44)<br>4 (9)<br>1 (2)<br>0 (0) |
| **3) Was the instructions clear?**<br>a. Yes<br>b. No | 45 (100)<br>0 (0) |
| **4) Having taken the sample yourself, would you have preferred to have completed it in a clinic?**<br>a. Yes<br>b. No | 4 (9)<br>41 (91) |
| **5) Did you require any support when taking the sample?**<br>a. Yes<br>b. No | 15 (33)<br>30 (67) |
| **6) If you needed to, were you able to contact a clinician for support?**<br>a. Yes<br>b. No<br>c. Not applicable | 25 (56)<br>1 (2)<br>19 (42) |

**Appendix Table 4:** ICA Feedback.

|  |  |
| --- | --- |
| Total | 45<br>n (%) |
| **1) How did you find using the ICA-Comp?**<br>a. Very Easy<br>b. Easy<br>c. Neither difficult nor easy<br>d. Difficult<br>e. Very Difficult | 3 (7%)<br>10 (22%)<br>13(29%)<br>9 (20%)<br>10 (22%) |
| **2) Were the instructions clear?**<br>a. Yes<br>b. No | 42(93%)<br>3 (7%) |
| **3) Did you require any support when completing the test?**<br>a. Yes<br>b. No | 14 (31%)<br>31 (69%) |

**Appendix Table 5:**
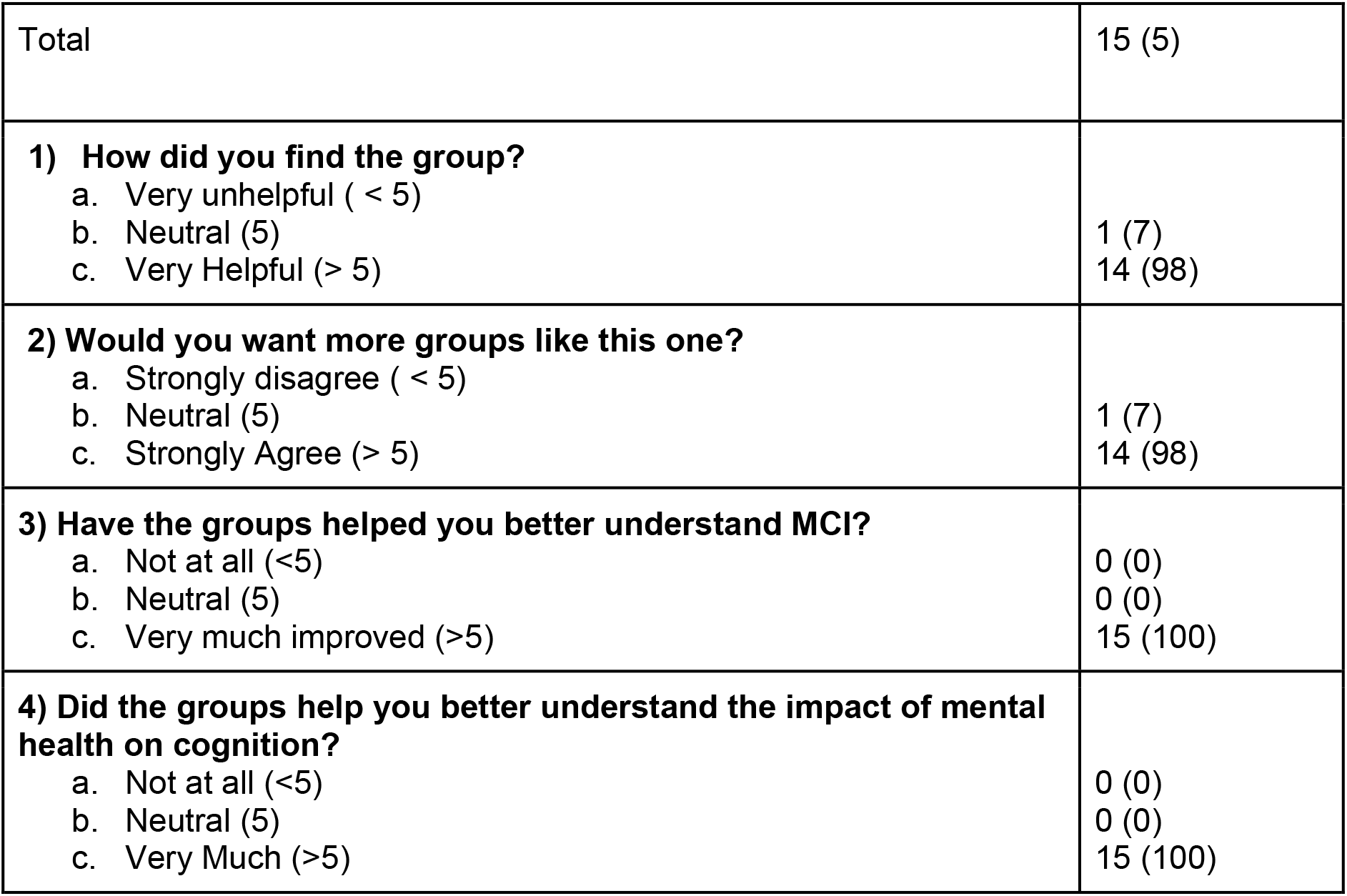
Cognitive Wellbeing Group Feedback, *with quantified response on a scale of 1-10 (with 10 reflecting a more positive response)*

## Notes

### Clinical Trial

NCT06379594

### Author Declarations

Research Ethics Committee (REC) 22/SC/0109 (South Central - Berkshire B, United Kingdom) gave ethical approval for this work.

